# Changes in health promoting behavior during COVID-19 physical distancing: Utilizing Wearable Technology to Examine Trends in Sleep, Activity, and Cardiovascular Indicators of Health

**DOI:** 10.1101/2020.06.07.20124685

**Authors:** Emily R Capodilupo, Dean J Miller

**Author notes:** Correspondence concerning this article should be addressed to: Dean J. Miller, Appleton Institute for Behavioral Science, Adelaide, South Australia, Australia.

## Abstract

The COVID-19 pandemic incited unprecedented restrictions on the behavior of society. The aims of this study were to quantify changes to sleep/wake behavior and exercise behavior, as well as changes in physiological markers of health during COVID-19 physical distancing. A retrospective analysis of 5,436 US-based subscribers to the WHOOP platform (mean age = 40.25 ± 11.33; 1,536 females, 3,900 males) was conducted covering the period from January 1st, 2020 through May 15th, 2020. This time period was separated into a 68-day baseline period and a 67-day physical distancing period. To provide context and allow for potential confounders (e.g., change of season), data were also extracted from the corresponding time periods in 2019. As compared to baseline, during physical distancing, all subjects fell asleep earlier (−0.25 hours), woke up later (0.48 hours), obtained more sleep (+0.35 hours) and reduced social jet lag (−0.21 hours). Contrasting sleep behavior was seen in 2019, with subjects falling asleep and waking up at a similar time (−0.01 hours; -0.05 hours), obtaining less sleep (−0.14 hours) and maintaining social jet lag (0.01 hours) in corresponding periods. Individuals exercised more intensely during physical distancing by increasing the time spent in high heart rate zones. In 2020, resting heart rate decreased (−0.9 beats per minute) and heart rate variability increased (+0.98 milliseconds) during physical distancing when compared to baseline. However, similar changes were seen in 2019, suggesting the variation may not be related to the introduction of physical distancing mandates. The findings suggest that changes in societal commitments (e.g., daily commute; working from home) during physical distancing may have resulted in changes to health-related behavior (i.e., increased exercise intensity and longer sleep duration). As the COVID-19 pandemic eases, maintenance of certain aspects of physical distancing (e.g., working from home) may allow for positive changes to sleep/wake and exercise behaviors.

## Introduction

Severe acute respiratory syndrome coronavirus 2 (SARS-CoV-2), the virus responsible for the COVID-19 pandemic [1], has caused significant social and economic disruptions and has altered the behavior of society. Millions of employees and business owners ceased working or shifted to working from home rather than commuting into offices. Physical distancing protocols put in place to reduce disease transmission also precluded previously common forms of group leisure and athletic activities. Exercise and sleep are two primary behaviors that may have been impacted by COVID-19 restrictions - with individuals confined to their homes and likely experiencing change to work (e.g., loss of employment; reduced commuting) and at-home commitments (e.g., childcare).

Changes in sleep/wake behavior can have meaningful implications on health. Insufficient sleep duration has been shown to correlate with impairments to cognitive performance [2], insulin sensitivity [3] and diabetes [4], cardiovascular disease [5], impairments to mental health [6], and reduced physical performance [7]. Increasing sleep quantity has been shown to counteract many of these effects; for example, increases in sleep quantity have been shown to drive changes in attention [8] and stress hormone expression [9] even within already healthy populations. Changes in sleep timing and the consistency of bedtimes and wake times also have profound effects on wellbeing, with benefits seen when a consistent sleep schedule is maintained [10]. These benefits are understood to be related to the circadian rhythm; shifting sleep timing decouples the circadian rhythm from the sleep/wake rhythm [11, 12], contributing to adverse health outcomes such as higher rates of cardiovascular disease [13] and breast cancer [14].

Physical distancing mandates brought with them temporary closure of gyms, banning of team sports, and cancelations of competitive seasons. For individuals that ordinarily exercise in these public settings, the way in which they obtain their daily exercise may have changed dramatically during the pandemic. Sudden changes in exercise modality, frequency and duration may have a variety of effects on fitness. For example, one study showed that women who participated in multiple exercise modalities had higher muscular endurance than women who participated in only one athletic activity [15]. Sudden changes in the exercise modalities performed by individuals may also result in changes to injury rates and [16] exercise intensity (e.g., transitioning from weight training to running). Therefore, if COVID-19 restrictions do provide more opportunity for exercise, increases in exercise frequency and intensity may result in beneficial cardiovascular adaptations [17]. Understanding how the distribution of time spent in each heart rate zone is perhaps as important as understanding how total exercise and exercise modality changed as a result of the COVID-19 pandemic. In this context, wearable technology can be leveraged to provide unique insight into health related behavior and subsequent health outcomes in large population cohorts.

The WHOOP strap (Whoop, Inc., Boston, MA, USA) is a commercially available wearable device that has been third-party validated to measure sleep, resting heart rate (RHR), heart rate variability (HRV), and respiratory rate [18]. Previous research has shown that metrics such as RHR [19] and HRV [20] are powerful indicators of cardiovascular health. Conditions brought forth by the pandemic create a natural testing ground in which to examine changes in health related behavior and whether these impact cardiovascular indicators of health. The primary aim of this study is to quantify the impact of physical distancing mandates on health related behaviors – specifically sleep timing and duration and exercise frequency, intensity, and modality. This study also explores the physical outcomes of these changes in behavior by examining markers of physical health, including sleep duration, RHR, and HRV.

## Materials and Methods

This study conducted a retrospective analysis of health-related behavior before and during COVID-19 physical distancing restrictions. A total of 5,436 WHOOP members based in the United States of America (mean age = 40.25 ± 11.33; 1,536 females, 3,900 males) were selected from the WHOOP subscriber-base. Inclusion criteria were (1) having recorded sleeps for at least 120 of the 135 (89%) days between January 1 and March 9 in 2019 and 2020, respectively; and (2) be between the ages of 18 and 80 on May 15th, 2020, when data was extracted for analysis. The study was approved by the Central Queensland University Human Research Ethics Committee. Data were collected with the written consent of individuals via WHOOP Inc’s terms of service.

To examine the acute changes in health related behavior, the period between January 1, 2020 and March 9th, 2020 was classified as a baseline period, while the period from March 10th, 2020 through May 15th, 2020 was classified as a physical distancing period. While no single date globally separates pre- and post-physical distancing, delineation of the data on March 9th is based on the rough timeline along which most users would have been subject to some level of physical distancing mandate or recommendation. During the week of March 9th, the World Health Organization officially classified COVID-19 as a pandemic [21] and a National State of Emergency was declared in the United States of America [22]. The May 15th physical distancing end date was chosen such that the physical distancing period analyzed would end before gradual easing physical distancing begins. Additionally, utilizing the chosen dates to represent physical distancing resulted in similar durations of baseline and physical distancing (68 and 67 days, respectively). To provide context for potential confounders (e.g., change in season), data were also extracted from the corresponding dates in 2019.

### Data extraction

The WHOOP strap (Boston, Massachusetts, USA) is a wearable device that collects continuous heart rate, 3-axis accelerometer, temperature, and 3-axis gyroscope data. In combination with the WHOOP mobile device app and cloud-based analytics platform, a series of algorithms evaluate duration and phases of sleep, RHR and HRV. Validation of sleep and cardiovascular measures gleaned from WHOOP against gold standard polysomnography support a low degree of bias and low precision errors [18, 23].

The WHOOP strap is capable of automatically detecting exercise and can automatically identify and classify 54 different modalities. Additionally, users of the WHOOP platform have the ability to manually log exercise that don’t meet the algorithm’s criteria for automatic detection.

The following variables were obtained from the WHOOP platform:

- Sleep opportunity duration: *total time dedicated to sleep each night. Measured primarily through automatic detection in the WHOOP app. Users are alerted each morning when their automatically detected sleep is ready for review and have the ability to edit the start and stop times should any errors occur*.
- Social jet lag: *the difference between sleep opportunity onset on weekends (Saturday and Sunday) and weekdays (Monday through Friday) resulting from misalignment between social and internal clocks [24]*.
- Sleep opportunity onset: *the time that each sleep opportunity was initiated relative to local time zone*.
- Sleep opportunity offset: *the time that each sleep opportunity ended relative to local time zone*
- Sleep duration (hours): *total amount of time spent asleep each night. Measured through automatic detection*.
- Exercise frequency: *daily percentage of individuals that recorded an exercise session*.
- Exercise type - *the sport or exercise type chosen for each exercise session. Labeled automatically by the app via a machine learning algorithm or manually by the WHOOP-user*.
- Exercise intensity: percentage of exercise *duration spent in each of the six heart rate zone*s -*Zone 1 = 0-50% heart rate reserve (HRR)* ; *zone 2 = 50-60% HRR, zone 3 = 60-70% HRR, zone 4 = 70-80% HRR, zone 5 = 80-90% HRR, zone 6 = 90-100% HRR. Measured automatically during the exercise session*.
- Resting heart rate (beats per minute; bpm) - *the mean value of heart beats per minute sampled during slow wave sleep. Automatically measured during slow wave sleep each night*.
- Heart rate variability (milliseconds; ms) - *the root-mean-square difference of successive heartbeat intervals sampled during slow wave sleep. Automatically measured during slow wave sleep each night*.

Throughout, dates are assigned to sleeps and exercise based on the local time zone’s date in which they end, for example, a sleep beginning in the final hours of January 1st and ending on the morning of January 2nd is attributed to January 2nd.

### Data Analysis

Independent non-parametric significance tests (Mann-Whitney U tests) were performed to examine differences in sleep and health related dependent variables between control and physical distancing periods in 2019 and 2020, respectively. Analyses for the 2020 time periods were also conducted at the cohort-level with data grouped by age and sex. An additional analysis examining changes between the 2020 control and physical distancing periods and corresponding changes in 2019 were also conducted. Where p<0.05, Bonferroni corrections were made to reduce the chance of obtaining a type 1 error. To assess the magnitude of the differences between these periods, Cohen’s *d* effect sizes and 95% confidence limits for effect size were also calculated. Effect sizes were interpreted as: <0.20 (trivial), 0.2 to 0.59 (small), 0.60 to 1.19 (moderate), 1.20 to 1.99 (large), and >2.0 (very large) [25].

## Results

### Sleep

A summary of sleep/wake behavior for age and gender cohorts are presented in Table 1. In 2020, average sleep opportunity duration was significantly longer during physical distancing when compared to baseline (*large effect size)*. During the corresponding time periods in 2019, sleep opportunity duration decreased (*small effect size)*. In 2020, average sleep opportunity duration was significantly longer during physical distancing when compared to baseline for 18-25 year-olds (*large effect size)*, 25-40 year-olds (*large effect size)*, 40-55 year-olds (*moderate effect size)*, 55-80 year-olds (*moderate effect size*), females (*large effect size*) and males (*moderate effect size*) (Table 1).

**Table 1.** Sleep/wake behavior as a function of time period.

| Variable | Time period |  | Outcomes |  |
| --- | --- | --- | --- | --- |
|  | Baseline<br>(M±SD) | Physical<br>distancing<br>(M±SD) | Difference<br>(PD-BL;<br>M±SEM) | Effect size<br>(95% CL) |
| **Sleep opportunity duration (h)** |  |  |  |  |
| 2019 | 7.70 ± 0.21 | 7.64 ± 0.19 | -0.05 ± 0.003* | -0.30 (-0.34, -0.30) |
| 2020 | 7.65 ± 0.20 | 7.89 ± 0.18 | 0.24 ± 0.003** | 1.26 (1.22, 1.30) |
| 18-25 year-olds | 7.83 ± 0.16 | 8.14 ± 0.16 | 0.31 ± 0.009** | 1.94 (1.77, 2.10) |
| 25-40 year-olds | 7.66 ± 0.19 | 7.93 ± 0.19 | 0.27 ± 0.004** | 1.42 (1.36, 1.48) |
| 40-55 year-olds | 7.56 ± 0.25 | 7.78 ± 0.21 | 0.21 ± 0.005** | 0.95 (0.89, 1.02) |
| 55-80 year-olds | 7.75 ± 0.19 | 7.90 ± 0.17 | 0.15 ± 0.007* | 0.83 (0.72, 0.94) |
| Females | 7.80 ± 0.2 | 8.07 ± 0.18 | 0.27 ± 0.005** | 1.42 (1.35, 1.49) |
| Males | 7.59 ± 0.2 | 7.81 ± 0.19 | 0.22 ± 0.003** | 1.13 (1.08, 1.17) |
| **Sleep onset time (hh:mm)** |  |  |  |  |
| 2019 | 22:59 ± 0:20 | 22:58 ± 00:15 | -00:01 ± 00:01 | -0.03 (-0.07, 0.00) |
| 2020 | 22:58 ± 0:19 | 23:13 ± 00:10 | 00:15 ± 00:01 | 1.02 (0.98, 1.06) |
| 18-25 year-olds | 23:31 ± 0:24 | 23:58 ± 00:13 | 00:26 ± 00:03** | 1.58 (1.42, 1.74) |
| 25-40 year-olds | 23:04 ± 0:23 | 23:21 ± 00:13 | 00:16 ± 00:01** | 1.13 (1.07, 1.18) |
| 40-55 year-olds | 22:49 ± 0:17 | 23:04 ± 00:09 | 00:14 ± 00:01** | 3.47 (3.39, 3.55) |
| 55-80 year-olds | 22:37 ± 0:13 | 22:47 ± 00:07 | 00:09 ± 00:01** | 3.18 (3.05, 3.31) |
| Females | 22:46 ± 0:19 | 23:04 ± 00:10 | 00:18 ± 00:01** | 3.10 (3.01, 3.18) |
| Males | 23:03 ± 0:19 | 23:17 ± 00:10 | 00:14 ± 00:01** | 1.10 (1.05, 1.14) |
| **Sleep offset time (hh:mm)** |  |  |  |  |
| 2019 | 06:43 ± 0:30 | 06:40 ± 00:27 | -00:03 ± 00:01 | -0.11 (-0.14, -0.07) |
| 2020 | 06:40 ± 0:29 | 07:09 ± 00:21 | 00:29 ± 00:01** | 1.15 (1.11, 1.18) |
| 18-25 year-olds | 07:24 ± 0:28 | 08:09 ± 00:19 | 00:45 ± 00:04** | 1.88 (1.71, 2.05) |
| 25-40 year-olds | 06:48 ± 0:31 | 07:20 ± 00:23 | 00:32 ± 00:01** | 1.17 (1.12, 1.23) |
| 40-55 year-olds | 06:26 ± 0:30 | 06:54 ± 00:21 | 00:27 ± 00:02** | 1.08 (1.02, 1.15) |
| 55-80 year-olds | 06:25 ± 0:21 | 06:44 ± 00:15 | 00:18 ± 00:02** | 1.04 (0.93, 1.15) |
| Females | 06:37 ± 0:28 | 07:11 ± 00:21 | 00:34 ± 00:02** | 1.37 (1.30, 1.45) |
| Males | 06:41 ± 0:30 | 07:09 ± 00:21 | 00:27 ± 00:01** | 1.08 (1.04, 1.13) |

Notes: BL = baseline; PD = physical distancing; SEM = standard error of the mean; \* = p-value
<0.05; \*\* = p-value <0.001.

In 2020, average sleep opportunity onset was 15 minutes later during physical distancing when compared to baseline (*moderate effect size)*. Average sleep opportunity onset was 1 minute earlier for the corresponding time periods in 2019 (*trivial effect size)*. In 2020, average sleep opportunity onset was significantly earlier during physical distancing when compared to baseline for 18-25 year-olds (*large effect size)*, 25-40 year-olds (*moderate effect size)*, 40-55 year-olds (*very large effect size)*, 55-80 year-olds (*very large effect size)*, females (*very large effect size)*, and males (*moderate effect size)* (Table 1). These data are presented in Fig 1.

**Fig 1.**
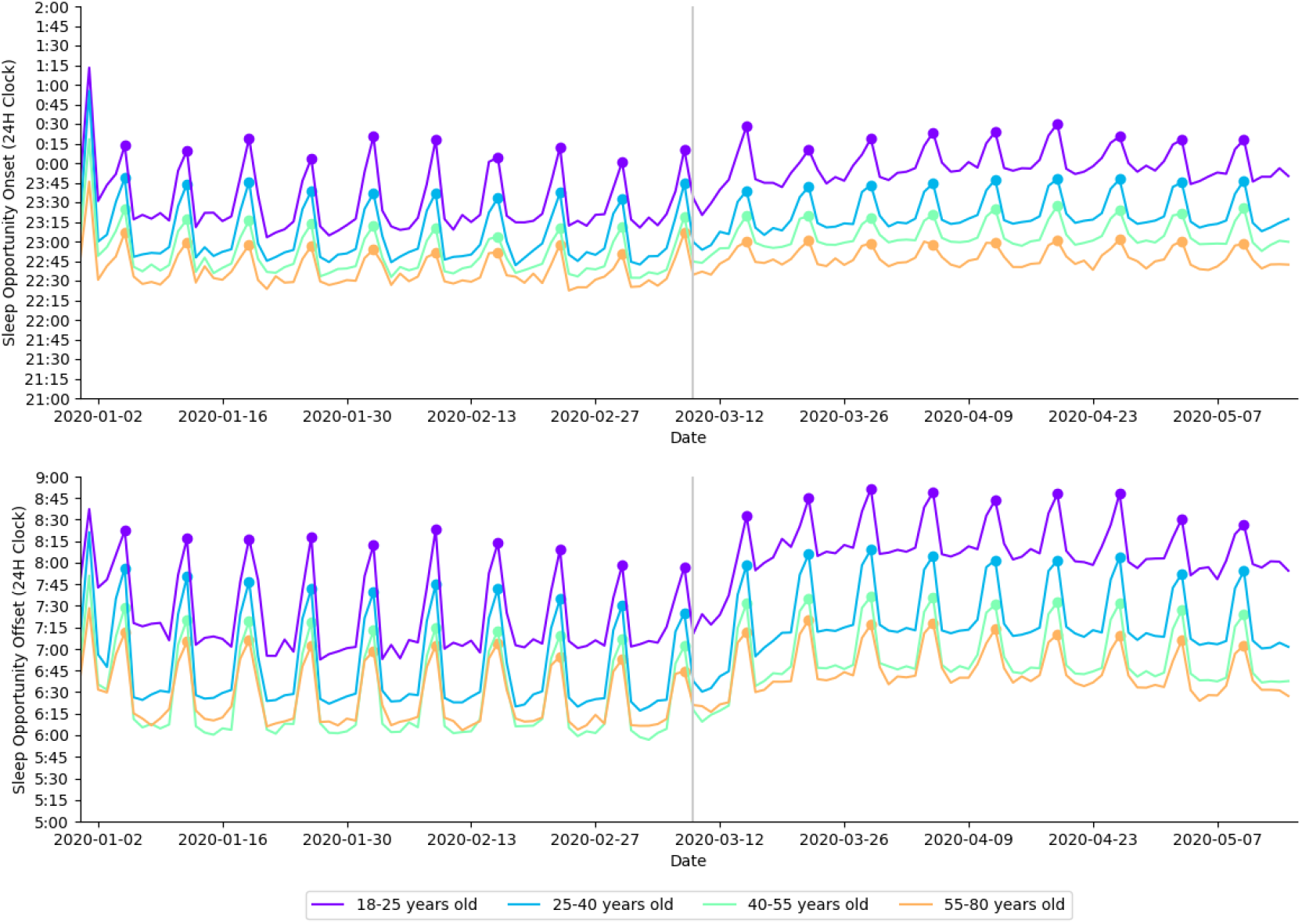
Daily average sleep opportunity onset time and sleep opportunity offset time in 2020 presented by age cohort. Each age cohort is plotted in a different color, indicted in the legend. Filled circles indicate Sundays; a gray vertical line separates baseline and physical distancing.

In 2020, average sleep opportunity offset was significantly later during physical distancing when compared to baseline (*moderate effect size)*. Average sleep opportunity offset was three minutes earlier for the corresponding time periods in 2019 (*trivial effect size)*. In 2020, average sleep opportunity offset was significantly later for 18-25 year-olds (*large effect size)*, 25-40 year-olds (*moderate effect size)*, 40-55 year-olds (*moderate effect size)*, 55-80 year-olds (*moderate effect size)*, females (*large effect size)*, and males (*moderate effect size)* (Table 1). These data are presented in Fig 1.

In 2020, average sleep duration was significantly longer during physical distancing when compared to baseline (*moderate effect size)*. Average sleep duration was significantly shorter for the corresponding time periods in 2019 (*very large effect size)*. In 2020, average sleep duration was significantly longer during physical distancing when compared to baseline for 18-25 year-olds (*very large effect size)*, 25-40 year-olds (*large effect size)*, 40-55 year-olds (*large effect size)*, 55-80 year-olds (*moderate effect size)*, 50-59 year-olds (*moderate effect size)*, females (*large effect size)*, and males (*large effect size)* (Table 2). These data are presented in Fig 2.

**Table 2.**
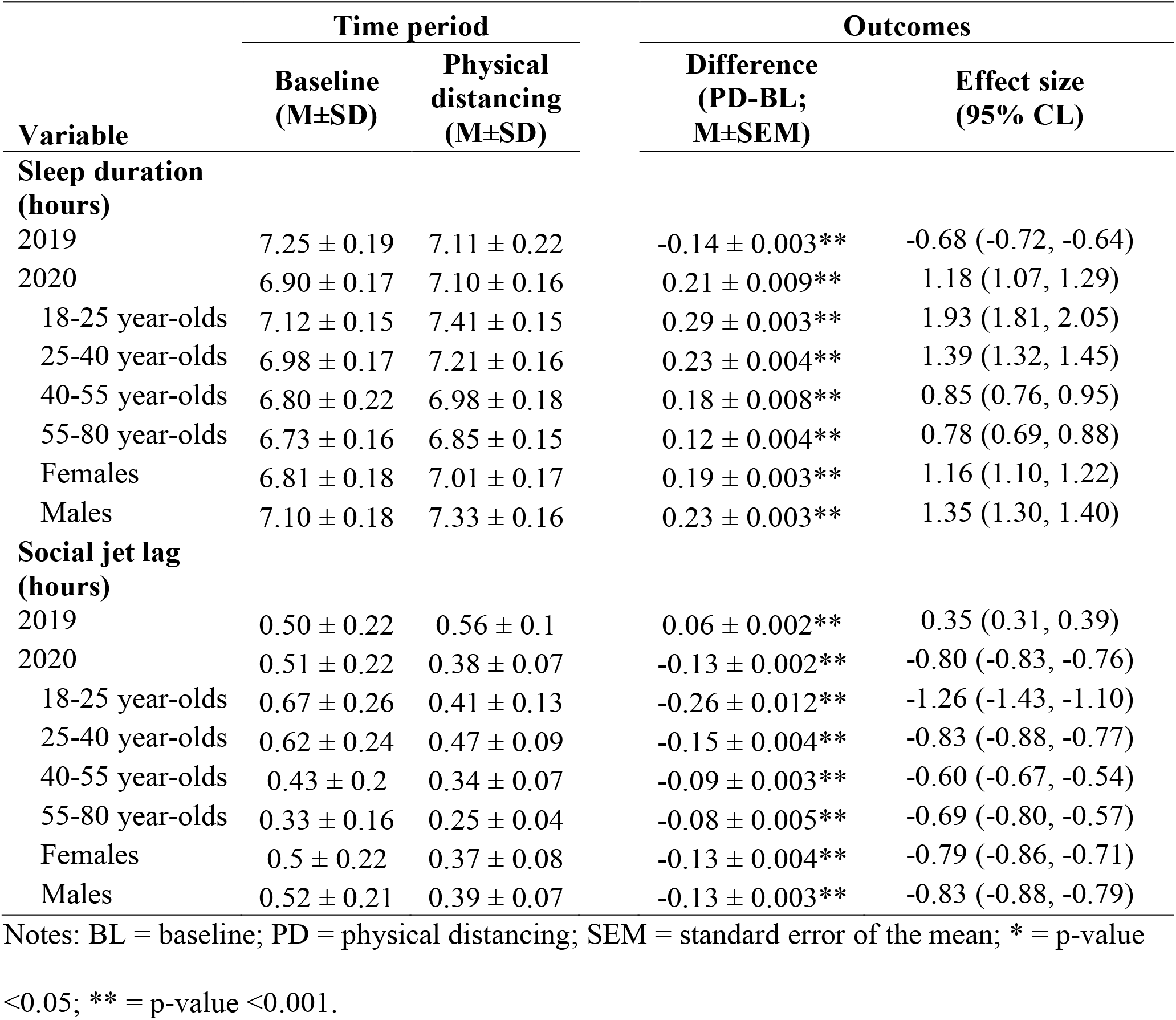
Objective sleep outcomes as a function of time period.

| Variable | Time period |  | Outcomes |  |
| --- | --- | --- | --- | --- |
|  | Baseline<br>(M±SD) | Physical<br>distancing<br>(M±SD) | Difference<br>(PD-BL;<br>M±SEM) | Effect size<br>(95% CL) |
| **Sleep duration**<br>**(hours)** |  |  |  |  |
| 2019 | 7.25 ± 0.19 | 7.11 ± 0.22 | -0.14 ± 0.003** | -0.68 (-0.72, -0.64) |
| 2020 | 6.90 ± 0.17 | 7.10 ± 0.16 | 0.21 ± 0.009** | 1.18 (1.07, 1.29) |
| 18-25 year-olds | 7.12 ± 0.15 | 7.41 ± 0.15 | 0.29 ± 0.003** | 1.93 (1.81, 2.05) |
| 25-40 year-olds | 6.98 ± 0.17 | 7.21 ± 0.16 | 0.23 ± 0.004** | 1.39 (1.32, 1.45) |
| 40-55 year-olds | 6.80 ± 0.22 | 6.98 ± 0.18 | 0.18 ± 0.008** | 0.85 (0.76, 0.95) |
| 55-80 year-olds | 6.73 ± 0.16 | 6.85 ± 0.15 | 0.12 ± 0.004** | 0.78 (0.69, 0.88) |
| Females | 6.81 ± 0.18 | 7.01 ± 0.17 | 0.19 ± 0.003** | 1.16 (1.10, 1.22) |
| Males | 7.10 ± 0.18 | 7.33 ± 0.16 | 0.23 ± 0.003** | 1.35 (1.30, 1.40) |
| **Social jet lag**<br>**(hours)** |  |  |  |  |
| 2019 | 0.50 ± 0.22 | 0.56 ± 0.1 | 0.06 ± 0.002** | 0.35 (0.31, 0.39) |
| 2020 | 0.51 ± 0.22 | 0.38 ± 0.07 | -0.13 ± 0.002** | -0.80 (-0.83, -0.76) |
| 18-25 year-olds | 0.67 ± 0.26 | 0.41 ± 0.13 | -0.26 ± 0.012** | -1.26 (-1.43, -1.10) |
| 25-40 year-olds | 0.62 ± 0.24 | 0.47 ± 0.09 | -0.15 ± 0.004** | -0.83 (-0.88, -0.77) |
| 40-55 year-olds | 0.43 ± 0.2 | 0.34 ± 0.07 | -0.09 ± 0.003** | -0.60 (-0.67, -0.54) |
| 55-80 year-olds | 0.33 ± 0.16 | 0.25 ± 0.04 | -0.08 ± 0.005** | -0.69 (-0.80, -0.57) |
| Females | 0.5 ± 0.22 | 0.37 ± 0.08 | -0.13 ± 0.004** | -0.79 (-0.86, -0.71) |
| Males | 0.52 ± 0.21 | 0.39 ± 0.07 | -0.13 ± 0.003** | -0.83 (-0.88, -0.79) |

Notes: BL = baseline; PD = physical distancing; SEM = standard error of the mean; \* = p-value
<0.05; \*\* = p-value <0.001.

**Fig 2.**
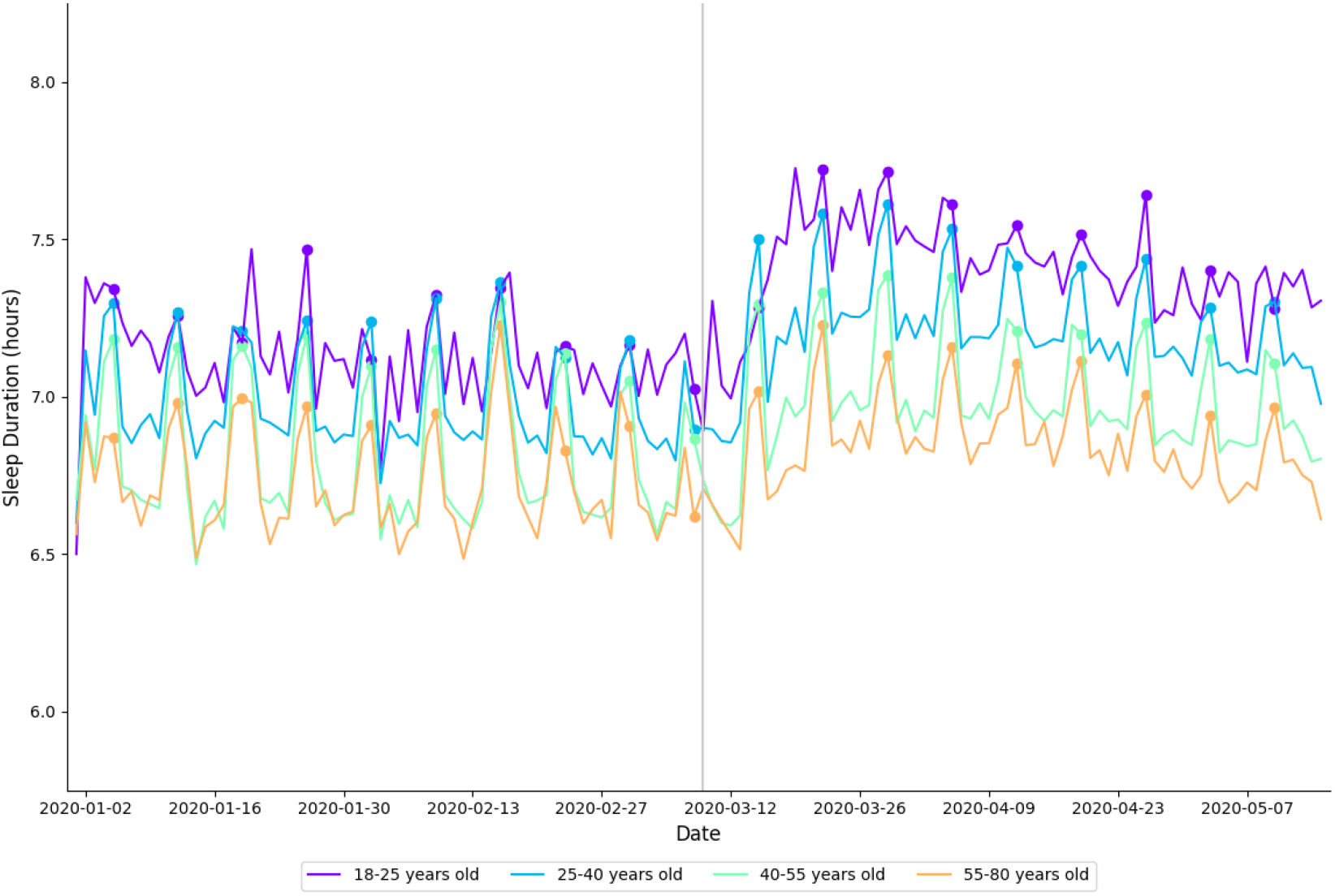
Daily average sleep duration in 2020 aggregated by age cohort. Each cohort is plotted in a different color, indicted in the legend. Filled circles indicate Sundays; a vertical gray line separates baseline and physical distancing.

In 2020, average social jet lag was significantly reduced during physical distancing when compared to baseline (*large effect size)*. Average social jet lag was significantly higher for the corresponding time periods in 2019 (*very large effect size)*. In 2020, average social jet lag was significantly reduced during physical distancing when compared to baseline for 18-25 year-olds (*very large effect size)*, 25-40 year-olds (*very large effect size)*, 40-55 year-olds (*very large effect size)*, 55-80 year-olds (*large effect size)*, females (*large effect size)*, and males (*large effect size)* (Table 2).

### Exercise

Summaries of exercise behavior for age and gender cohorts are presented in Table 3 and Fig 3. In 2020, there was no difference in exercise frequency during physical distancing when compared to baseline (*small effect size)*. There was no difference in exercise intensity for the corresponding time periods in 2019 (*trivial effect size)*. In 2020, exercise frequency did not differ between baseline and physical distancing for 18-25 year-olds (*small effect size)*, 25-40 year-olds (*small effect size)*, 40-55 year-olds (*moderate effect size)* and males (*small effect size)*. Exercise frequency increased during physical distancing when compared to baseline for 55-80 year-olds (*moderate effect size)* and females (*small effect size)* (Table 3). These data are presented in Fig 3.

**Table 3.** Daily percentage of users that engaged in exercise (i.e., exercise frequency) as a function of time period.

| Cohort | Exercise frequency (%) |  | Outcomes |  |
| --- | --- | --- | --- | --- |
|  | Baseline (M±SD) | Physical distancing (M±SD) | Difference (PD-BL; M±SEM) | Effect size (95% CL) |
| 2019 | 66.98 ± 4.14 | 67.0 ± 4.67 | 0.02 ± 0.06 | 0.00 (-0.03, 0.04) |
| 2020 | 59.9 ± 4.04 | 61.91 ± 4.36 | 2.01 ± 0.06** | 0.48 (0.44, 0.52) |
| **Age (years)** |  |  |  |  |
| 18-25 year-olds | 65.62 ± 10.44 | 62.83 ± 8.22 | -2.79 ± 0.05** | -0.30 (-0.45, -0.14) |
| 25-40 year-olds | 60.09 ± 5.01 | 61.7 ± 4.77 | 1.61 ± 0.10* | -0.80 (-0.86, -0.75) |
| 40-55 year-olds | 58.72 ± 3.31 | 61.85 ± 4.33 | 3.13 ± 0.09** | 0.46 (0.39, 0.52) |
| 55-80 year-olds | 59.27 ± 2.75 | 62.17 ± 5.14 | 2.89 ± 0.16** | 0.84 (0.73, 0.95) |
| **Sex** |  |  |  |  |
| Females | 63.29 ± 5.49 | 65.43 ± 5.16 | 2.14 ± 0.14* | 1.16 (1.08, 1.23) |
| Males | 58.53 ± 3.61 | 60.45 ± 4.21 | 1.92 ± 0.06** | -0.72 (-0.77, -0.68) |

Notes: BL = baseline; PD = physical distancing; SEM = standard error of the mean; \* = p-value
<0.05; \*\* = p-value <0.001.

**Fig 3.**
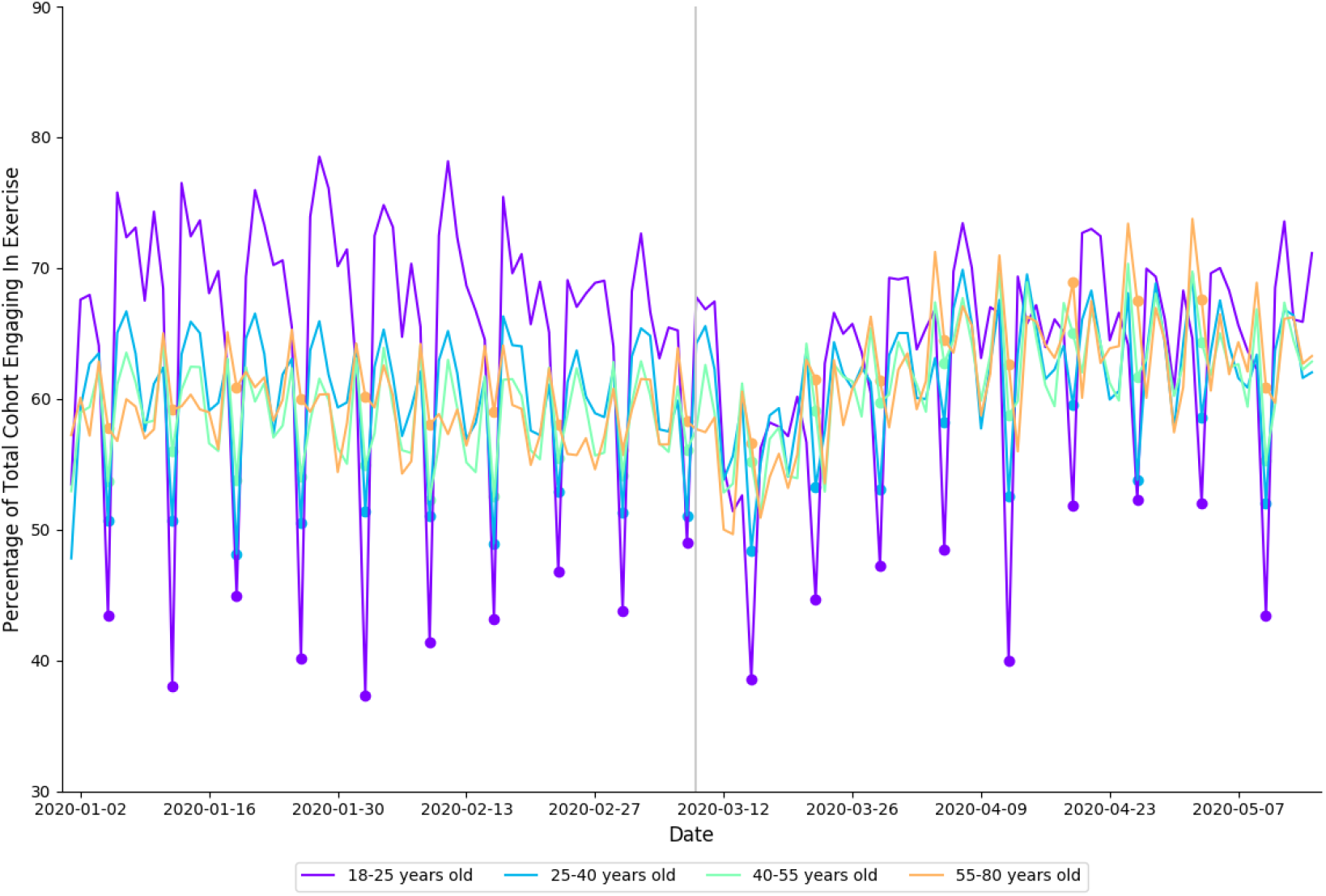
Percentage of each age cohort in 2020 that logged an exercise session, by date. Each cohort is shown in a different color, indicated in the legend. Filled circles indicate Sundays, a vertical gray line separates baseline from physical distancing.

In 2020, individuals spent less time in HR zone 1 (*small effect size)*, HR zone 2 (*moderate effect size)* and HR zone 3 (*large effect size);* and spent more time in HR zone 4 (*small effect size)*, HR zone 5 (*moderate effect size)*, and HR zone 6 (*moderate effect size)* during physical distancing compared to baseline (Table 4). For the corresponding periods in 2019, individuals spent less time in HR zone 1, and more time in HR zone 5 and HR zone 6. These data are presented in Fig 4.

**Table 4.** Distribution of exercise intensity as a function of time period.

| HR Zone | Time period |  | Outcomes |  |
| --- | --- | --- | --- | --- |
|  | Baseline<br>(M±SD) | Physical<br>distancing<br>(M±SD) | Difference<br>(PD-BL;<br>M±SEM) | Effect size<br>(95% CL) |
| 2019 |  |  |  |  |
| Zone 1 | 11.94 ± 0.51 | 10.94 ± 0.93 | -1.0 ± 0.01** | -1.33 (-1.37, -1.29) |
| Zone 2 | 21.43 ± 1.08 | 20.82 ± 1.19 | -0.61 ± 0.015* | -0.54 (-0.57, -0.50) |
| Zone 3 | 28.84 ± 0.57 | 29.25 ± 0.51 | 0.41 ± 0.007** | 0.76 (0.72, 0.80) |
| Zone 4 | 24.28 ± 0.86 | 24.8 ± 1.06 | 0.52 ± 0.013* | 0.54 (0.50, 0.58) |
| Zone 5 | 12.22 ± 0.95 | 12.77 ± 1.14 | 0.55 ± 0.014* | 0.52 (0.49, 0.56) |
| Zone 6 | 1.30 ± 0.15 | 1.42 ± 0.19 | 0.12 ± 0.002** | 0.70 (0.66, 0.74) |
| 2020 |  |  |  |  |
| Zone 1 | 11.66 ± 0.8 | 12.09 ± 1.18 | 0.43 ± 0.014* | 0.43 (0.39, 0.46) |
| Zone 2 | 20.32 ± 1.17 | 19.54 ± 0.91 | -0.78 ± 0.014** | -0.74 (-0.78, -0.71) |
| Zone 3 | 27.73 ± 0.62 | 26.58 ± 0.54 | -1.15 ± 0.008** | -1.98 (-2.02, -1.94) |
| Zone 4 | 25.54 ± 1.12 | 25.98 ± 1.19 | 0.44 ± 0.016* | 0.38 (0.34, 0.42) |
| Zone 5 | 13.42 ± 1.11 | 14.33 ± 0.69 | 0.91 ± 0.013** | 0.98 (0.95, 1.02) |
| Zone 6 | 1.33 ± 0.16 | 1.48 ± 0.12 | 0.15 ± 0.002** | 1.06 (1.02, 1.10) |

Notes: BL = baseline; PD = physical distancing; SEM = standard error of the mean; \* = p-value
<0.05; \*\* = p-value <0.001.

**Fig 4.**
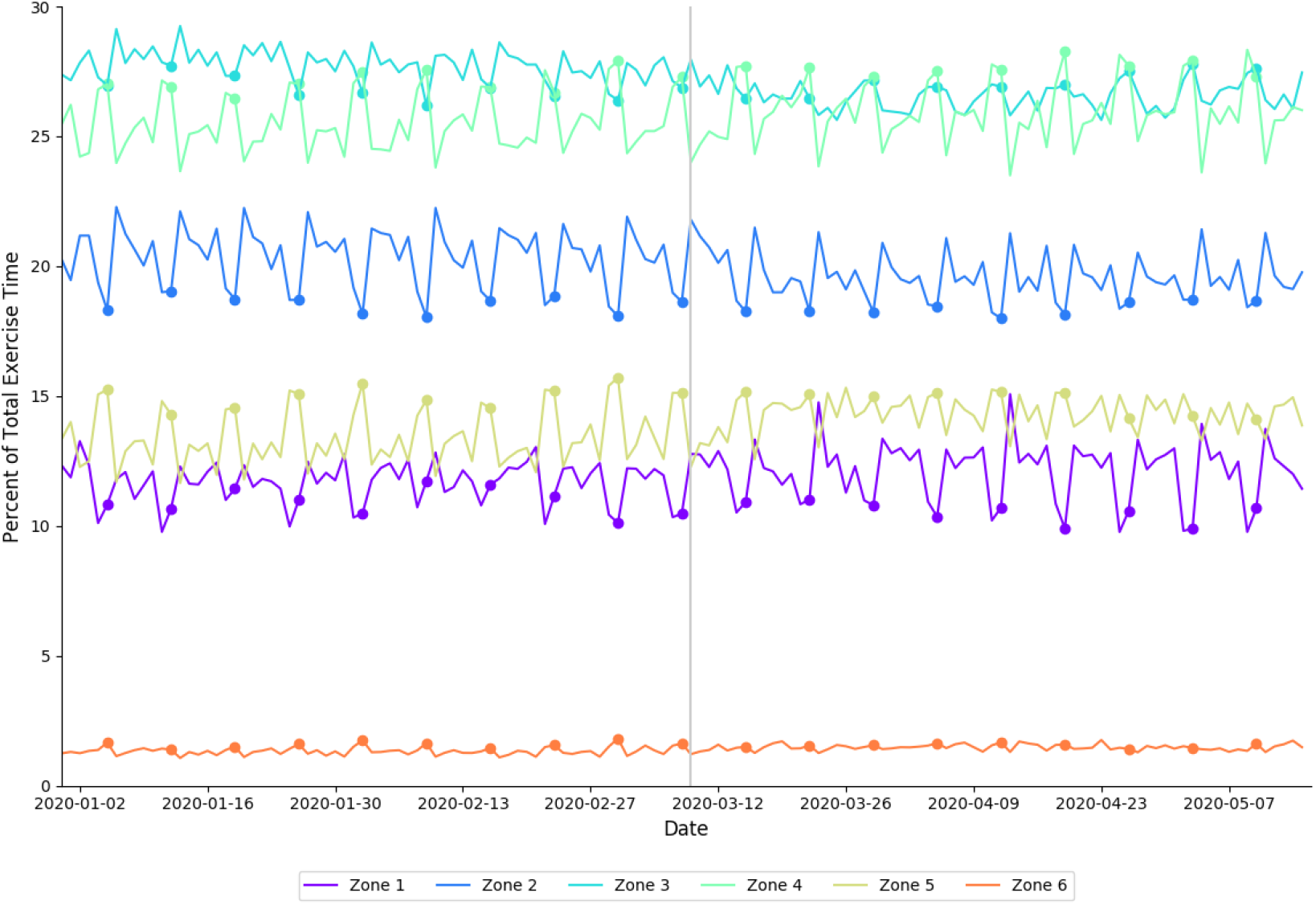
Distribution of proportion of total exercise time spent in each heart rate zone in 2020, arranged by date. Each heart rate zone is shown in a different color, indicated by the legend. Filled circles indicate Sundays; the vertical gray line delineates between baseline and physical distancing.

### Cardiovascular indicators of health

A summary of cardiovascular outcomes for age and gender cohorts are presented in Table 5. In 2020, average RHR was lower during physical distancing when compared to baseline (*large effect size)*. For the corresponding periods in 2019, average RHR was lower (moderate effect size). In 2020, average RHR was lower during physical distancing when compared to baseline for 18-25 year-olds (*moderate effect size)*, 25-40 year-olds (*moderate effect size)*, 40-55 year-olds (*large effect size)*, 55-80 year-olds (*large effect size)*, females (*large effect size)*, and males (*large effect size)* (Table 5). These data are presented in Fig 5.

**Table 5.**
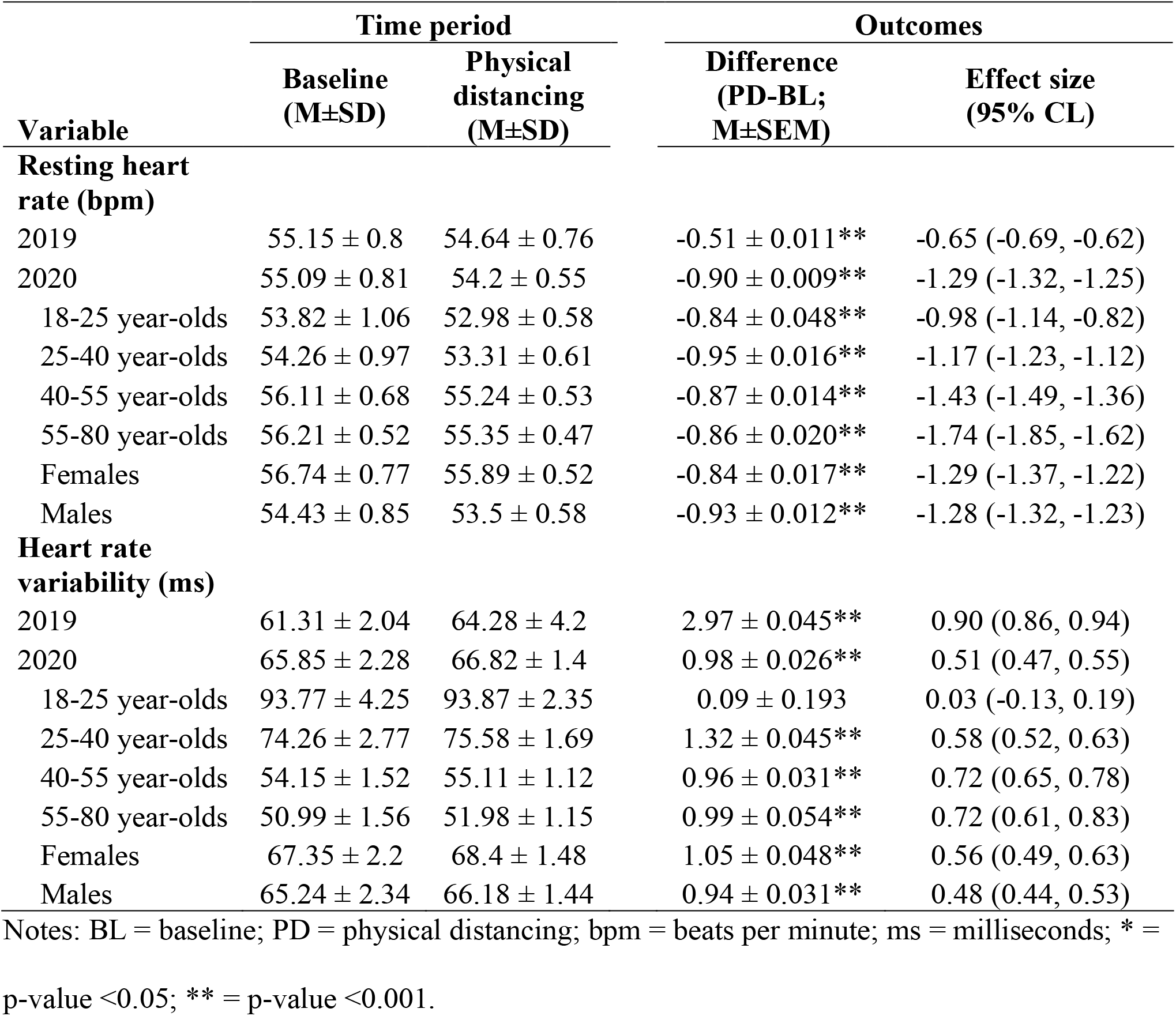
Cardiovascular outcomes as a function of time period.

| Variable | Time period |  | Outcomes |  |
| --- | --- | --- | --- | --- |
|  | Baseline<br>(M±SD) | Physical<br>distancing<br>(M±SD) | Difference<br>(PD-BL;<br>M±SEM) | Effect size<br>(95% CL) |
| **Resting heart rate (bpm)** |  |  |  |  |
| 2019 | 55.15 ± 0.8 | 54.64 ± 0.76 | -0.51 ± 0.011** | -0.65 (-0.69, -0.62) |
| 2020 | 55.09 ± 0.81 | 54.2 ± 0.55 | -0.90 ± 0.009** | -1.29 (-1.32, -1.25) |
| 18-25 year-olds | 53.82 ± 1.06 | 52.98 ± 0.58 | -0.84 ± 0.048** | -0.98 (-1.14, -0.82) |
| 25-40 year-olds | 54.26 ± 0.97 | 53.31 ± 0.61 | -0.95 ± 0.016** | -1.17 (-1.23, -1.12) |
| 40-55 year-olds | 56.11 ± 0.68 | 55.24 ± 0.53 | -0.87 ± 0.014** | -1.43 (-1.49, -1.36) |
| 55-80 year-olds | 56.21 ± 0.52 | 55.35 ± 0.47 | -0.86 ± 0.020** | -1.74 (-1.85, -1.62) |
| Females | 56.74 ± 0.77 | 55.89 ± 0.52 | -0.84 ± 0.017** | -1.29 (-1.37, -1.22) |
| Males | 54.43 ± 0.85 | 53.5 ± 0.58 | -0.93 ± 0.012** | -1.28 (-1.32, -1.23) |
| **Heart rate variability (ms)** |  |  |  |  |
| 2019 | 61.31 ± 2.04 | 64.28 ± 4.2 | 2.97 ± 0.045** | 0.90 (0.86, 0.94) |
| 2020 | 65.85 ± 2.28 | 66.82 ± 1.4 | 0.98 ± 0.026** | 0.51 (0.47, 0.55) |
| 18-25 year-olds | 93.77 ± 4.25 | 93.87 ± 2.35 | 0.09 ± 0.193 | 0.03 (-0.13, 0.19) |
| 25-40 year-olds | 74.26 ± 2.77 | 75.58 ± 1.69 | 1.32 ± 0.045** | 0.58 (0.52, 0.63) |
| 40-55 year-olds | 54.15 ± 1.52 | 55.11 ± 1.12 | 0.96 ± 0.031** | 0.72 (0.65, 0.78) |
| 55-80 year-olds | 50.99 ± 1.56 | 51.98 ± 1.15 | 0.99 ± 0.054** | 0.72 (0.61, 0.83) |
| Females | 67.35 ± 2.2 | 68.4 ± 1.48 | 1.05 ± 0.048** | 0.56 (0.49, 0.63) |
| Males | 65.24 ± 2.34 | 66.18 ± 1.44 | 0.94 ± 0.031** | 0.48 (0.44, 0.53) |

Notes: BL = baseline; PD = physical distancing; bpm = beats per minute; ms = milliseconds; \* =
p-value <0.05; \*\* = p-value <0.001.

**Fig 5.**
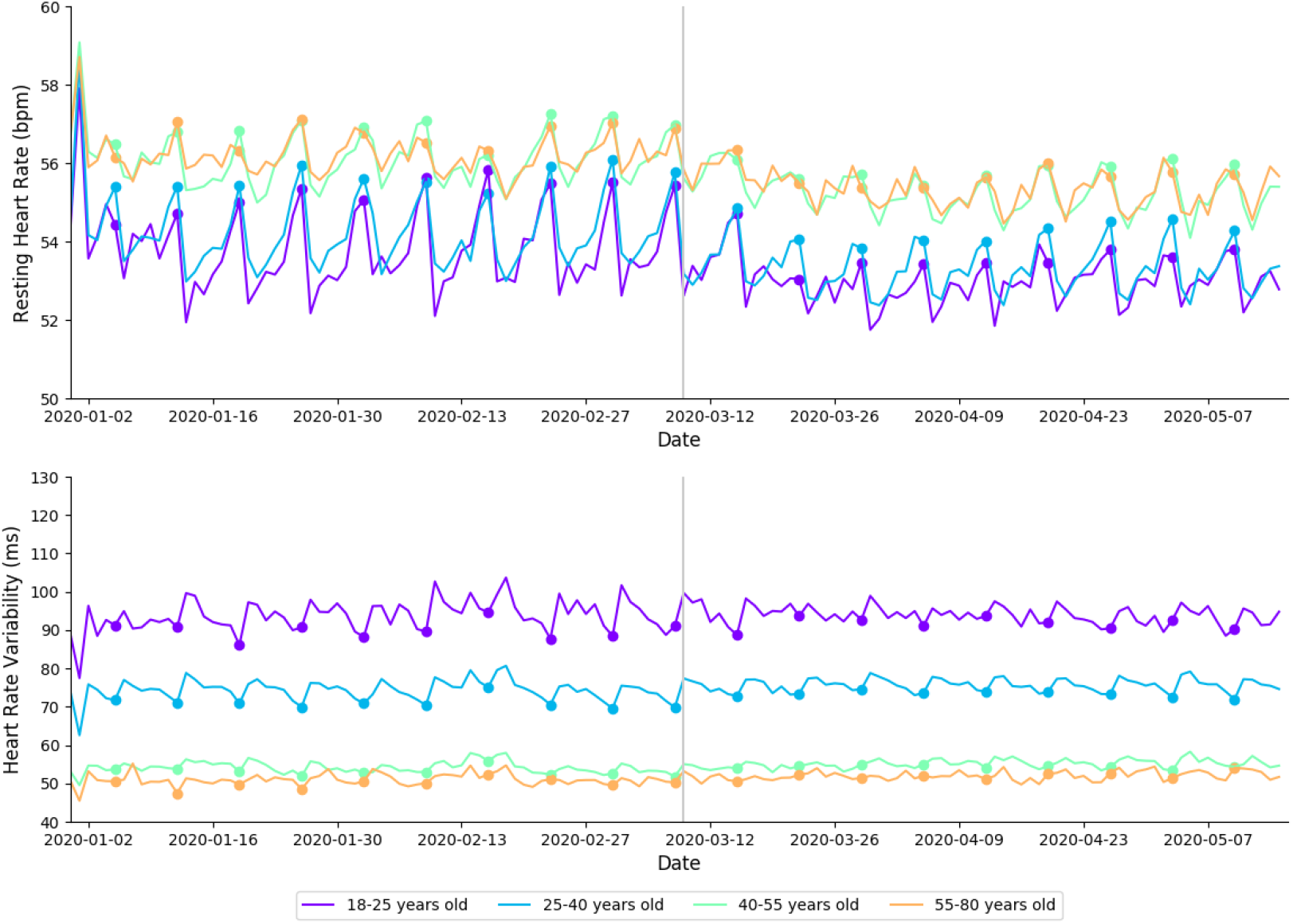
Changes in RHR (top) and HRV (bottom) in 2020 arranged by date and age cohort. Each cohort is shown with a differently colored line as indicated in the legends. bpm = beats per minute; ms = milliseconds. Filled circles indicate Sundays; a gray vertical line separates baseline from physical distancing.

In 2020, average HRV was higher during physical distancing when compared to baseline (*large effect size)*. Average HRV increased over the corresponding time periods in 2019 (*moderate effect size*). In 2020, average HRV was higher during physical distancing when compared to baseline for 18-25 year-olds (*moderate effect size)*, 25-40 year-olds (*moderate effect size)*, 40-55 year-olds (*large effect size)*, 55-80 year-olds (*large effect size)*, females (*large effect size)*, and males (*large effect size)* (Table 5). These data are presented in Fig 5.

### Comparison of 2019 and 2020

A comparison of changes to sleep and exercise variables during designated time periods (i.e., BL-PD) between 2019 and 2020 are presented in Table 6. Subjects significantly increased sleep opportunity duration (*large effect size)*, delayed sleep offset time (*moderate effect size)*, had an earlier sleep offset time (*moderate effect* size), increased sleep duration (*large effect size)* and reduced social jet lag (*moderate effect size)* during PD in 2020 when compared to the shift in the corresponding time periods in 2019 (Table 6). Subjects decreased exercise frequency *(very large effect size)* during PD in 2020 when compared to the changes seen in the corresponding time periods in 2019. For exercise intensity, subjects significantly decreased the proportion of time spent in zones 2 (*trivial effect size)*, 3 (*very large effect size)* and 6 (*trivial effect sizes)*, and significantly increased the proportion of time spent in zones 1 (*large effect size)*, 4 (*trivial effect size)*, and 5 (*small effect size)* during PD in 2020 when compared to the changes in the corresponding time periods in 2019. Subjects reduced RHR (*small effect size)* and increased HRV (*moderate effect size*) during PD in 2020 when compared to the changes seen in the corresponding time periods in 2019.

**Table 6.** Differences in sleep and exercise related variables as a function of year.

| Variable | Year |  | Outcomes |  |
| --- | --- | --- | --- | --- |
|  | 2019<br>(BL-PD;<br>M±SD) | 2020<br>(BL-PD;<br>M±SD) | Difference<br>(2020-2019;<br>M±SEM) | Effect size<br>(95% CL) |
| Sleep opportunity duration (h) | -0.05 ± 0.20 | 0.24 ± 0.19 | 0.29 ± 0.003 | 1.49 (1.45, 1.53) |
| Sleep onset time (hh:mm) | 00:00 ± 0:18 | 00:15 ± 0:15 | 00:15 ± 00:00 | 0.91 (0.87, 0.94) |
| Sleep offset time (hh:mm) | -00:03 ± 0:28 | -00:29 ± 0:25 | -00:26 ± 00:00 | 0.98 (0.94, 1.02) |
| Sleep duration (h) | -0.14 ± 0.20 | 0.21 ± 0.17 | 0.35 ± 0.003 | 1.89 (1.85, 1.93) |
| Social jet lag (h) | 0.00 ± 0.20 | -0.20 ± 0.20 | -0.2 ± 0.003 | -1.00 (-1.04, -0.96) |
| Exercise frequency (%) | 0.02 ± 0.04 | 2.01 ± 0.04 | 1.99 ± 0.001 | 49.75 (49.38, 50.12) |
| Exercise intensity |  |  |  |  |
| Zone 1 (%) | -1.0 ± 0.75 | 0.43 ± 1.0 | 1.43 ± 0.012 | 1.62 (1.58, 1.66) |
| Zone 2 (%) | -0.61 ± 1.13 | -0.78 ± 1.04 | -0.17 ± 0.015 | -0.16 (-0.19, -0.12) |
| Zone 3 (%) | 0.41 ± 0.54 | -1.15 ± 0.58 | -1.56 ± 0.007 | -2.78 (-2.83, -2.74) |
| Zone 4 (%) | 0.52 ± 0.96 | 0.44 ± 1.15 | -0.08 ± 0.014 | -0.08 (-0.11, -0.04) |
| Zone 5 (%) | 0.55 ± 1.04 | 0.91 ± 0.92 | 0.36 ± 0.013 | 0.37 (0.33, 0.40) |
| Zone 6 (%) | 0.12 ± 0.17 | 0.15 ± 0.14 | 0.03 ± 0.002 | 0.19 (0.16, 0.23) |
| RHR (bpm) | -0.51 ± 0.78 | -0.90 ± 0.69 | -0.39 ± 0.010 | -0.53 (-0.57, -0.49) |
| HRV (ms) | 2.97 ± 3.27 | 0.98 ± 1.87 | -1.99 ± 0.036 | -0.75 (-0.79, -0.71) |

Notes: BL = baseline; PD = physical distancing; bpm = beats per minute; ms = milliseconds; \* =
p-value <0.05; \*\* = p-value <0.001.

## Discussion

The aim of this study was to detail changes in health related behavior and outcomes associated with the introduction of physical distancing mandates during the COVID-19 pandemic. A retrospective analysis of sleep and activity patterns was conducted covering January 1, 2020 through May 15, 2020 including 5,436 US-based WHOOP members. Given that the timeframes in the study included changes of season and daylight saving, a corresponding dataset from 2019 was included to provide additional context. Findings were divided into sleep, activity, and cardiovascular indicators of health and are discussed below.

### Sleep

The primary sleep related findings were (1) sleep onset shifted later during physical distancing; (2) average sleep duration increased in all age and gender cohorts during physical distancing and (3) social jet lag – i.e., weekday-to-weekend variability of sleep opportunity onset and offset – decreased for all gender and age cohorts during physical distancing.

The findings of this study suggest that the implementation of physical distancing mandates may have impacted the sleep/wake behavior of this population. Along with increased sleep opportunity duration during physical distancing, an apparent shift towards a later sleep onset and offset was evident. It appears that subjects were able to obtain more sleep by delaying sleep offset. It can be suggested that this behavioral change may be due to reduced morning commitments during physical distancing (e.g., commute). The shifts seen in 2019 support this finding, with the same subjects exhibiting earlier wake times (Table 1) and less total sleep in the corresponding periods (Table 2). A similar delay in sleep timing during COVID-19 restrictions have been reported in multiple survey-based studies [26, 27, 28]. In regard to changes among age cohorts, 18-25 year-olds experienced the largest delay their sleep offset time. This finding reflects the established differences in chronobiology across age cohorts - younger individuals tend to have a late chronotype while older individuals skew towards an earlier chronotype [29]. Given that school and collegiate based commitments likely reduced during physical distancing – the data suggests that teenagers and young adults returned to a more chronobiologically suitable sleep period (i.e., go to bed late, wake up late).

In addition to exhibiting differences in the magnitude of average sleep timing shifts, Fig 1 highlights a significant reduction in the social jet lag during physical distancing. In contrast, a significant increase in social jet lag was seen in 2019. It is likely that the reduction in social jet lag seen in 2020 were due to restrictions on social or extra-curricular activities during physical distancing. As may be expected, this was most prominent in the younger cohorts, with larger reductions seen when compared to older cohorts (Table 2). In support of our findings, a recent global survey of 11,431 adults from 40 countries found that social jet lag reduced by approximately 30 minutes during physical distancing restrictions [27].

Average nightly sleep duration in 2020 was significantly higher during physical distancing than during baseline for all age cohorts (Fig 1). This finding is in contrast to the reduction in sleep duration seen in 2019 (Table 6) – suggesting that this change was likely due to the shifts in sleep/wake behavior experienced during physical distancing. Once again, larger changes were seen in the younger age cohorts (i.e., 18-25 year old’s). These findings suggest that physical distancing may have alleviated societal factors (e.g., work or academic commitments, commuting) that restrict sleep opportunity, allowing individuals to revert towards biological sleep/wake preferences. Practical applications can be made across all age cohorts; however, the data highlight important changes in behavior amongst the 18-25 year-old cohort. As mentioned above, individuals from this age cohort typically have late chronotypes (i.e., their circadian drive for sleep initiates later in the night). However, they are often expected to fulfil academic or sport related commitments early in the morning - decreasing opportunity for sleep. Prior literature has demonstrated subjective and objective benefits associated with even minor extension to average sleep duration when sustained over time; a 2018 study by Lo et al. [30] showed that 10 minutes of extended sleep duration in secondary school students correlated with improved alertness and mental health. These findings show that despite their typically late chronotype, 18-25 year-olds will shift their sleep offset later if given the opportunity. In addition to improvements in cognition, sleep extensions have been shown to correspond with statistically significant reductions in systolic blood pressure [31].

### Exercise

The primary exercise findings were that (1) a higher proportion of individuals performed exercise during physical distancing; (2) individuals increased exercise intensity during physical distancing, and (3) individuals changed the types of athletic activities they were performing during physical distancing. The changes in activities observed are consistent with adherence to bans on gatherings of more than 10 people and closures of athletic facilities such as fitness clubs and gyms. Coincidentally, athletic activities that are compliant to physical distancing appear to be those that also require high cardiovascular load (i.e., running, cycling). This is supported by a decrease in weightlifting and increases in activities like running (Fig 6). This increase may be due to the minimal equipment needs (e.g., pair of sneakers for running), and that individuals are likely to prioritize outdoor activities whilst confined to their homes for much of each day. Previous studies suggest that engagement in new exercise modalities may increase exercise intensity [32], improve athletic performance and improve cardiovascular health [33, 34]. Therefore, alterations to training stimulus (i.e., exercise modality), and the associated physiological adaptations, may be an unexpected side effect of physical distancing restrictions.

**Fig 6.**
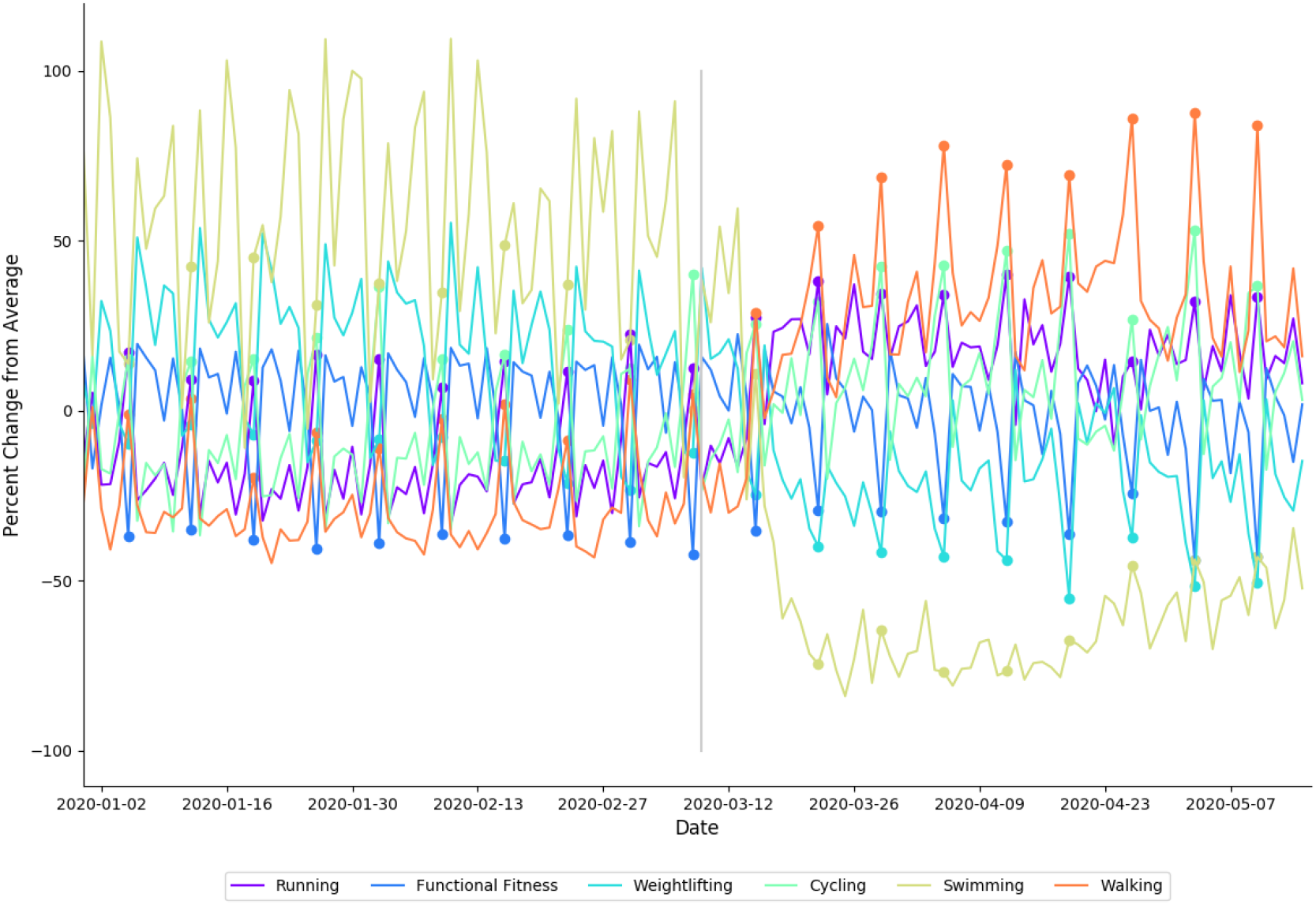
Percentage of change from average for the six most popular exercise modalities. Each modality is shown with a differently colored line as indicated in the legends. Filled circles indicate Sundays; a gray vertical line separates baseline from physical distancing.

In addition to changes in exercise modalities performed, our data show that individuals exercised more intensely during physical distancing (Fig 4). In 2020, the highest three heart rate zones (collectively, 70-100% HRR) occupy a larger proportion of total exercise time during physical distancing than they did during baseline (Table 4). Similar increases were seen in Zones 4, 5 and 6 in 2019, however there was a significantly larger shift seen in Zone 5 during 2020 (Table 6). In 2020, distributions in relative time spent in heart rate zones 2 through 5 show a cyclical pattern with a 7-day period that is more pronounced during baseline than physical distancing; relatively more time is spent in zones 4, 5, and 6 on weekends, while more time is spent in zones 2 and 3 on weekdays (Fig 4). The differentiation between weekend and weekday heart rate zone distributions is less pronounced in all heart rate zones during physical distancing than it was during baseline. This suggests that individuals were performing more intense exercise on weekdays during physical distancing when compared to baseline. An increase in time spent in these higher heart rate zones may mean an increase in anaerobic training, which has been previously demonstrated to reduce RHR and improve endurance [35]. The present study highlights changes in exercise modality and increases in exercise intensity in this subset of WHOOP users during physical distancing restrictions. Irrespective of physical distancing mandates, it is physiologically sound to suggest that such behaviors, if sustained, should result in improved health outcomes [36].

### Cardiovascular Indicators of health

The main finding regarding cardiovascular indicators of health was that average RHR decreased and HRV increased during physical distancing (**Fig 3**). Both outcomes represent an improvement in cardiovascular health – however it should be noted that similar shifts were seen in 2019. In particular, the increase in HRV seen in 2019 was larger than the increase seen in 2020 (Table 6). While the mechanisms behind these differences are unclear, HRV is known to be sensitive to changes in a range of behavioral factors (e.g., exposure to novel training stimuli, alcohol consumption; overtraining). A possible explanation for the smaller increase seen in 2020 may be that increased exercise intensity during physical distancing increased the contribution of the sympathetic nervous system – therefore resulting smaller acute increases in HRV.

For RHR, there was a larger reduction in 2020 compared to 2019, suggesting that changes to sleep and exercise behaviors during physical distancing may have had positive benefits on cardiovascular health. Analysis of the baseline period shows expected moderate differences across age cohorts in which younger individuals show signs of greater cardiovascular fitness than older cohorts. While it is well documented that HRV declines with age [37, 38], previous studies have demonstrated no age-related increase in RHR. For example, Kostis et al [39] found no relationship between age and RHR; this apparent discrepancy may be related to the small effect size and the relatively small datasets of previous studies. Therefore, a novel finding of the present study is that older cohorts presented with progressively higher RHR.

### Limitations and boundary conditions

Interpretations of the present results must be made with consideration to the boundary conditions. The WHOOP user demographic is likely to be more health conscious, have a higher socioeconomic status and are more likely to have the ability to work from home. The findings of this study may not be representative of global changes in behavior due to all individuals being in the United States of America. While it is reasonable to suggest that individuals from this demographic might have decreased professional commitments during physical distancing, it is also possible that exercise opportunity may decrease due to childcare or other at-home commitments. No data were collected regarding the mental health of participants and the severity and/or adherence to physical distancing mandates among the subjects. The date chosen to distinguish between the respective periods was an estimated timeline along which most users would have been subject to some level of physical distancing mandate or recommendation. The high number of statistical comparisons (i.e., multiple testing) and large sample size may have increased the likelihood of significant findings in the analyses. However, Bonferroni corrections were made to reduce the likelihood of type 1 errors due to multiple testing and Cohen’s d effect sizes were used to assess the magnitude of each effect.

## Conclusions

The aim of this study was to report on the sleep and exercise behavioral changes associated with COVID-19 related physical distancing mandates. The findings suggest that physical distancing mandates may have resulted in changes to sleep and exercise patterns, which may have consequences on the health and wellbeing of this demographic. While improved sleep and exercise patterns appear to be the mechanism for positive health-related behavior change during physical distancing, it is unclear which specific barriers were limiting these activities prior to physical distancing. It is reasonable to assume that decreases in business hours-based commitments (e.g., commuting) has previously impacted health related behavior (i.e., exercise and sleep). Therefore, in the context of a post-pandemic society, increased flexibility in how business, academia and other professional endeavors are conducted (i.e., ability to work from home) may allow individuals more opportunity to engage in health enhancing behaviors. Future research will be required as countries begin to reopen to investigate which behaviors are maintained, how these behaviors impact mental health, and if cohorts return to their previous behaviors.

## Data Availability

Data cannot be shared publicly due to user privacy.

